# Assessing the risk of vaccine-driven virulence evolution in SARS-CoV-2

**DOI:** 10.1101/2020.12.01.20241836

**Authors:** Ian F. Miller, C. Jessica E. Metcalf

**Affiliations:** Department of Ecology and Evolutionary Biology, Princeton University; Princeton School of Public and International Affairs

## Abstract

How might COVID-19 vaccines alter selection for increased SARS-CoV-2 virulence, or lethality? Framing current evidence surrounding SARS-CoV-2 biology and COVID-19 vaccines in the context of evolutionary theory indicates that prospects for virulence evolution remain uncertain. However, differential effects of vaccinal immunity on transmission and disease severity between respiratory compartments could select for increased virulence. To bound expectations for this outcome, we analyze an evo-epidemiological model. Synthesizing model predictions with vaccine efficacy data, we conclude that while vaccine driven virulence evolution remains a theoretical risk, it is unlikely to threaten prospects for herd immunity in immunized populations. Given that this event would nevertheless impact unvaccinated populations, virulence should be monitored to facilitate swift mitigation efforts.

**Significance statement:** Vaccines can provide personal and population level protection against infectious disease, but these benefits can exert strong selective pressures on pathogens. Virulence, or lethality, is one pathogen trait that can evolve in response to vaccination. We investigated whether COVID-19 vaccines could select for increased SARS-CoV-2 virulence by reviewing current evidence about vaccine efficacy and SARS-CoV-2 biology in the context of evolutionary theory, and subsequently analyzing a mathematical model. Our findings indicate that while vaccine-driven virulence evolution in SARS-CoV-2 is a theoretical risk, the consequences of this event would be limited for vaccinated populations. However, virulence evolution should be monitored, as the ramifications of a more virulent strain spreading into an under-vaccinated population would be more severe.

## Introduction

Since its emergence in late 2019, SARS-CoV-2 has spread globally, resulting in over 150 million cases of its associated disease, COVID-19, and over 3 million deaths (1). Beyond the morbidity and mortality associated with the pandemic, nations and sub-national states heavily affected by COVID-19 have experienced catastrophic economic collapse (2), critical pauses in educational services, and pervasive psychological damage. Generating herd immunity through vaccination has been consistently identified as the only acceptable course of action for mitigating the pandemic and allowing society to begin the recovery process. The race for SARS-CoV-2 vaccines has proceeded at an unprecedented pace. Following promising results from phase 3 trials (3–6) several vaccines have been authorized for emergency use. While clinical trials provide the evidence necessary to confirm the ability of vaccines to safely prevent disease in individuals, potential population level and longer-term consequences of vaccine introduction, particularly viral evolution, should also be considered. Potential negative outcomes include vaccine escape via antigenic evolution, which would result in a decrease or loss of vaccine efficacy (7), and the evolution of increased virulence, which could result in more severe health outcomes and a higher infection fatality ratio amongst unvaccinated individuals. Here, we focus on the latter outcome, and assess the potential for vaccines to drive the evolution of SARS-CoV-2 virulence. First, we review the theory surrounding the evolution of virulence evolution as it relates to SARS-CoV-2. Next, we consider the effects of COVID-19 vaccines in this theoretical framework, evaluating current evidence and assessing the potential for selection for increased virulence using an evo-epidemiological model. Finally, we synthesize our findings and generate recommendations for minimizing this risk, a question of growing importance in the happy circumstance of ever greater deployment of vaccines globally.

### The theory of virulence evolution

The evolution of disease associated damage or mortality, termed ‘quantitative virulence’, has intrigued scientists for decades. In the absence of any associated costs, pathogens are expected to evolutionarily maximize both the rate at which they transmit from infected hosts, and the duration of time during which a host is infectious. Because mortality (and in some cases severe symptoms) curtails opportunities for transmission, virulence alone is never adaptive for pathogens, and is at best selectively neutral (e.g. when host mortality occurs after the host infectious period). However, for many pathogens, host damage either can enhance transmission (e.g. by inducing coughing) or is an unavoidable consequence of or requirement for transmission (e.g. cellular damage resulting from viral replication). Thus, selection for increased virulence can occur as a result of selection for increased transmission (8–10), and indeed, only as a result of selection for increased transmission.

From this evolutionary theory, it is evident that two conditions must be met for virulence evolution to occur. First, increased transmission must be biologically feasible, and must be determined by the genetics of the pathogen (e.g., it must be heritable). Second, evolutionary increases in transmission must lead to increases in virulence (e.g. virulence must be an unavoidable consequence of transmission). These two conditions determine whether virulence evolution is possible, but they are not informative about the limits to virulence evolution, i.e., how destructive the pathogen has the potential to become, in terms of host mortality.

There are two major sets of factors that limit virulence evolution. The first involves limits to transmission evolution. Because virulence can only be selected for in association with transmission, any biologic, metabolic, or other limits to transmission evolution (such as limits to the rate of protein synthesis or viral assembly) necessarily constrain virulence evolution (11, 12). The second set of factors limiting virulence evolution encompasses the effects of host mortality on transmission rate. If neither symptoms nor disease-associated death shorten the duration of transmission, then virulence does not reduce pathogen fitness and its evolution is not constrained (13). Conversely, if symptoms or disease can truncate the infectious period, then the potential for a trade-off between virulence and transmission that constrains the evolution of these two traits emerges.

To put this formally, for many pathogens, incremental increases in transmission are hypothesized to bear increasing costs of damage, leading to a positive, saturating relationship between mortality (or severity) rate and the maximum attainable transmission rate. This relationship -- the canonical ‘virulence transmission tradeoff’-- has the effect of limiting the evolution of virulence (Fig. 1). The saturating nature of this trade-off curve makes an intermediate degree of virulence evolutionarily optimal, as the benefits of increased transmission are balanced against the costs of host death (or other severe outcomes truncating transmission). If the trade-off curve does not saturate, then any decreases in transmission time due to increased virulence are more than compensated for by increases in transmission rate, and the evolution of virulence is not constrained. It is important to note that the trade-off curve bounds the set of possible virulence and transmission strategies. Evolutionary changes can occur on the interior of this set with unconstrained increases and/or decreases in both virulence and transmission, and as such the presence or shape of a trade-off may not be apparent until transmission is maximized for a given virulence strategy and evolution begins to ‘trace the curve.’ Direct evidence for the existence of virulence-transmission trade-offs is limited, but suggestive evidence has been found in many systems (10).

**Figure 1:**
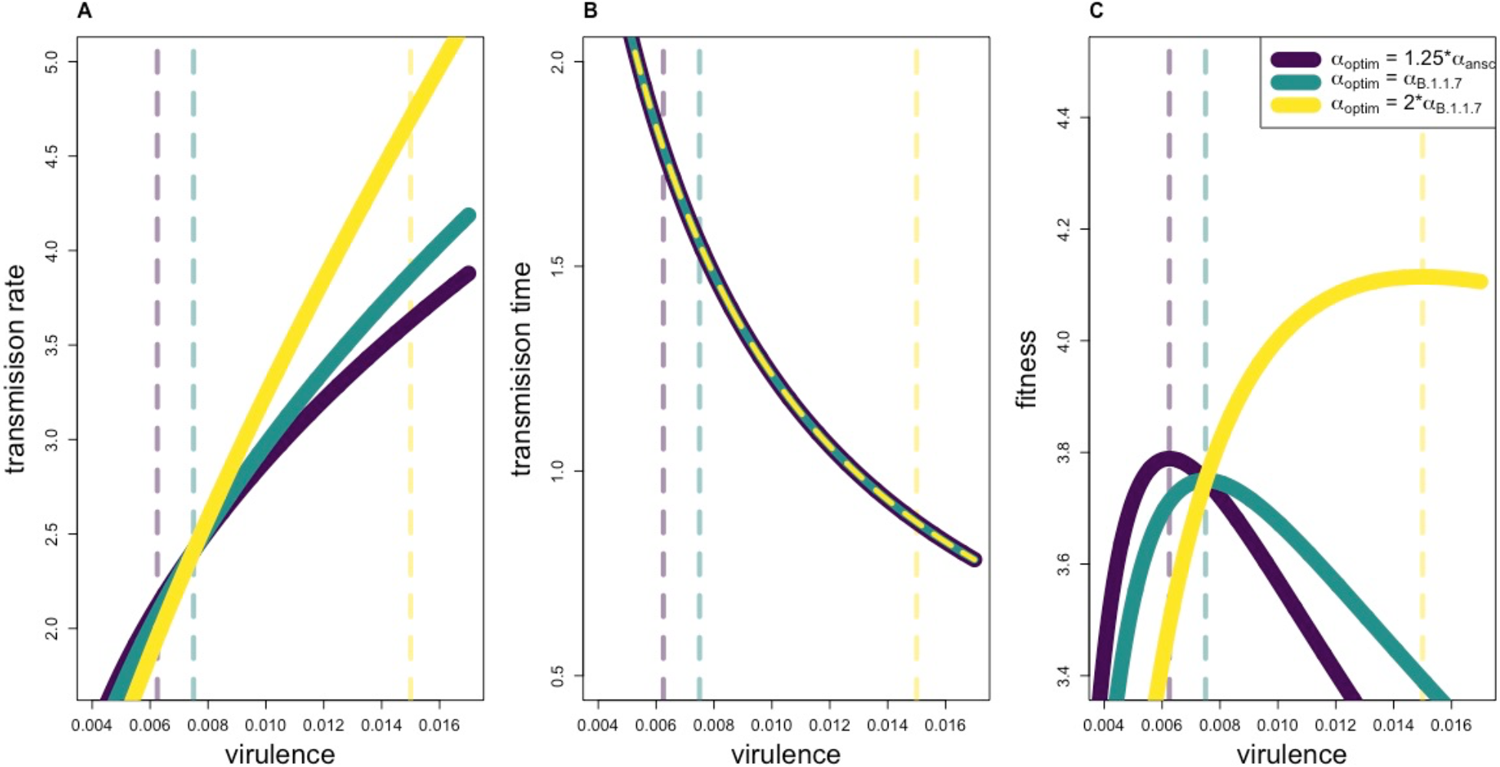
Virulence-transmission trade-offs. **(A)** different possible shapes for a virulence-transmission trade-off. The curves define the maximum transmission rate attainable for a given virulence strategy (interpretable as e.g., the infection fatality ratio). The region under each curve represents the set of all possible combinations of transmission rate and virulence for that trade-off shape. **(B)** The relationship between virulence and transmission time, assuming *p* = 66.666 (see methods). **(C)** The relationship between virulence and fitness (in a completely susceptible population, equivalent to R_0_) for different trade-off shapes. Yellow, teal, and purple curves in panels A-C correspond to *α*_*optium*_ = 1.25 * *α*_*ansc*,_ *α*_*B*.1.1.7_ and 2* *α*_*B*.1.1.7_ respectively, and reflect relationships unaffected by immunity (see methods). Vertical dashed lines show the optimum virulence strategies corresponding to each curve.

These theoretical frameworks can be used to assess whether the evolution of increased SARS-CoV-2 virulence is possible and to identify factors limiting the extent to which this could occur.

### Evaluating potential for virulence evolution in SARS-CoV-2

The evidence surrounding differences in transmission and virulence between the ancestral SARS-CoV-2 strain and emergent variants sheds light on whether the two conditions necessary for SARS-CoV-2 virulence evolution are met. While only a few variants have been definitively linked to increased transmission, current evidence gives a clear indication that the first condition (the feasibility and heritability of increased transmission rate) is satisfied. The *B*.*1*.*1*.*7* strain (i.e. VOC 202012/01, 20I/501Y.V) has been identified as being 43-82% transmissible than other circulating genotypes (14), although factors like founder-effects might overinflate this estimate of transmissibility. Likewise, the *D614G* mutation is hypothesized to be associated with increased transmission due to the rapid increase in its frequency (15, 16), enhanced replication in vitro (16, 17) and increased viral titers in vivo (18). While current evidence clearly supports the first condition (potential for increased transmission), support for the second condition (virulence being a consequence of increased transmission) is mixed. Preliminary analyses indicate that the *B*.*1*.*1*.*7* variant is associated with an approximately 55% increase in infection fatality rate (19), but clinical data suggests that the *D614G* mutation is not associated with a change in the severity of disease (20). Together, these lines of evidence indicate that future evolutionary increases in SARS-CoV-2 virulence are possible, but not certain.

Given that the evolution of increased SARS-CoV-2 virulence is possible, could either biological limits to transmission or a virulence-transmission trade-off constrain how virulent SARS-CoV-2 could become? To date, no limits to the evolution of SARS-CoV-2 transmission rate have been described. However, despite extremely large viral population sizes, only a few evolutionary increases in transmission have been detected, but this pattern should not necessarily be viewed as indicative of future trends. Turning to the potential for a virulence-transmission trade-off to limit the evolution of virulence, several threads of epidemiological and clinical evidence indicate that disease associated mortality does not significantly decrease transmission time in SARS-CoV-2 infection: The infectious period for COVID-19 infection is known to start before symptom onset (21), the majority of infected individuals experience only mild symptoms (22), and death usually occurs several weeks after initial infection (23). These patterns don’t preclude the existence of a virulence-transmission trade-off, but they do suggest that if such a trade-off does exist, virulence likely limits transmission time through a mechanism other than death. This could occur if increased virulence is associated with increased symptom frequency (i.e. a lower asymptomatic rate, shown not to be the case for B.1.1.7 (24)), more severe initial symptoms for those who are symptomatic, or faster symptom onset. Due to widespread public knowledge of SARS-CoV-2 circulation and non-pharmaceutical interventions such as case isolation (which has been shown to decrease transmission time as evidenced by a shortening of the serial interval (25)), individuals infected with a more virulent variant would presumably more frequently and/or more rapidly modify their behavior to reduce onwards transmission than individuals infected with a less virulent strain. Overall, while a virulence-transmission trade-off could emerge from the relationship between transmission rate and mortality rate, or these potential links between severity, symptoms, and transmission time, current data are insufficient to conclusively determine the existence of such a trade-off, its shape, or where current the SARS-CoV-2 virulence and transmission rates fall on the curve or within the set of strategies that it bounds.

### COVID-19 vaccines and virulence evolution

Current evidence indicates that the evolution of increased SARS-CoV-2 virulence is possible, as transmission and virulence may be linked, but the constraints determining the extent to which virulence might increase are not well resolved. Considering this prospect is nevertheless important, because vaccinal immunity can determine the directionality of selection for increased virulence in certain circumstances. If the evolution of transmission is unconstrained, then selection always favors increased transmission (and thus increased virulence if the two are associated) regardless of the effects of immunity, which can only modify the strength of selection. However, if virulence evolution is constrained by a virulence-transmission trade-off, vaccines might have the potential to determine the strength and directionally of selection for increased virulence by removing or reducing the costs of virulence to the pathogen. In this case, theory predicts that immunity which reduces disease but not transmission (26), or reduces disease to a greater extent than transmission (27), can drive the evolution of increased virulence. This points to a potential risk for certain vaccines to drive the evolution of more virulent pathogens. While there is clear evidence for some aspects of vaccine driven pathogen evolution (e.g. vaccine escape) in several human diseases, to date there have been no increases in virulence definitively linked to vaccine use in humans (28). However, several pieces of evidence highlight the existence of this risk: Evolutionary increases in virulence driven by vaccination have been observed in a mouse model of malaria (29) and in an oncogenic herpes virus infecting commercially raised chickens (30).

Vaccinal immunity to SARS-CoV-2 could reduce disease to a greater extent than transmission, potentially opening the door for virulence evolution, if its impacts on transmission and mortality/disease severity are not equal. The contrasting consequences of SARS-CoV-2 infection in the upper respiratory tract (URT) and lower respiratory tract (LRT) provide a framing mechanism for separating the effects of immunity on these factors. Evidence to date indicates that protection in the LRT could reduce disease severity, potentially reducing the costs of increased transmission. Protection in the URT, the primary location of infection colonization, might lead to a reduction in infection risk, potentially even to the level of sterilizing immunity, negating any gains in transmission rate the virus might be able to acquire via increased virulence (31). The partitioning of virulence and transmission effects between respiratory tract compartments is known to exist for other respiratory diseases such as influenza (32, 33). In SARS-CoV-2, the mechanisms underlying the separation of virulence and transmission effects between the LRT and URT are increasingly resolved. The pattern of higher viral infectivity in the URT compared to the LRT reflects a decreasing gradient of angiotensin-converting enzyme 2 (ACE2, the receptor protein utilized by SARS-CoV-2 for cellular entry) expression from the URT to the LRT (34).

Given this structure, if vaccines have differential effects in the LRT and URT, then they may have differential effects on transmission and virulence, opening the way to their driving virulence evolution. Preliminary evidence suggests that protective effects of SARS-CoV-2 vaccines might indeed differ between respiratory compartments. Non-human primate challenge studies investigating the efficacy of early COVID-19 vaccine candidates found that vaccinal immunity reduced viral replication in the LRT to a greater extent than in the URT (31, 35). The patterns of lower URT than LRT protection for these candidate vaccines may be rooted in the type of immune responses they induce. All are delivered intramuscularly, which generally and predominantly stimulates the production of IgG antibodies. The LRT system is primarily protected by these IgG antibodies, while the URT is primarily protected by IgA antibodies involved in mucosal immunity (31). Other tissues where SARS-CoV-2 infection can cause acute injury (36) are protected by IgG antibodies, indicating immunological protection against damage in the lower respiratory and the rest of the body might be correlated.

While the scale of vaccine distribution indicates that vaccines will be a critical factor shaping the landscape of immunity that drives SARS-CoV-2 evolution in many parts of the world, naturally acquired immunity could also contribute to selective pressures. Evidence suggests that similarly to SARS-CoV-2 vaccines, natural immunity provides greater LRT protection than URT protection, although the differential might not be as large. In macaques re-challenged with SARS-CoV-2, immunity eliminated or significantly reduced viral replication in the LRT, and reduced replication in the URT, but to a lesser extent (37). Consistent with this pattern, a longitudinal study of healthcare workers in the UK found that 0/1246 individuals with anti-spike protein IgG antibodies and 89/ 11052 seronegative individuals became symptomatically infected, indicating that natural immunity provides robust protection against disease (38). The same study also found that natural immunity provides substantial but incomplete immunity against infection, as the incidence of asymptomatic infection was observed to be roughly four times higher in the seronegative group compared to the seropositive group (38). Natural infection stimulates the production of both IgG and IgA antibodies (31), which suggests that natural infection could generate more balanced URT and LRT protection than vaccination, making the evolution of increased virulence a less likely outcome of selection imposed by natural immunity alone.

### Bounding the effects of COVID-19 vaccines on selection for increased virulence

The unprecedented scale and speed at which SARS-CoV-2 vaccines are likely to be deployed point to considerable value in bounding expectations on selection for increased viral virulence under different vaccine characteristics. The uncertainty surrounding the plausibility of and limits to SARS-CoV-2 virulence evolution complicates predictions, but it is possible to identify conditions associated with a pessimistic scenario, in which vaccination has the most potential to determine both the directionality and the strength of selection for increased virulence. These conditions include the heritability of and thus evolutionary potential for increased transmission, a positive association between transmission and virulence, no biological limits to transmission, diminishing gains in transmission rate with increasing virulence, and increases in the removal rate of infectious individuals proportional to increases in virulence. These last two conditions constitute a virulence-transmission trade-off, and while they intentionally do not represent a specific mechanism, they are broadly consistent with a variety of ways in which a trade-off might emerge (e.g. faster symptom onset, more frequent isolation of symptomatic individuals).

We develop a general theoretical framework to explore the degree to which different combinations of vaccinal protection in the URT and LRT (represented by parameters *r*_*U,V*_ and *r*_*L,V*_) might shape selection for a variant with increased virulence (relative to B.1.1.7, modeled as 1.5\**α*_*B*.1.1.7_) under the conditions associated with this ‘pessimistic’ scenario (Fig. 2). Our approach is focused on evaluating patterns of selection for a moderate increase in virulence in the short-term, and as such our results are not informative about the ‘endpoint’ of virulence evolution in the long term or patterns of selection for extremely virulent variants. To accommodate our mechanism-free assumption of a virulence-transmission trade-off, we assume susceptible-infectious-recovered epidemiological dynamics. While this model formulation does not explicitly incorporate details of COVID-19 epidemiology (e.g. pre-symptomatic transmission), its general form broadly captures all relevant dynamics. To explore how the strength and direction of selection for increased virulence changes with assumptions about where the virulence of *B*.*1*.*1*.*7* falls on the virulence-transmission trade-off curve, we replicate our analyses across a range of assumptions about how optimal virulence, *α*_*optium*_, compares to the virulence associated with *B*.*1*.*1*.*7* (*α*_*obs*_), which is the most-fit variant detected to date. Our analysis is motivated by SARS-CoV-2, but separation of contributions to virulence and transmission from different compartments (e.g., in URT and LRT) is likely relevant for other viruses.

**Figure 2:**
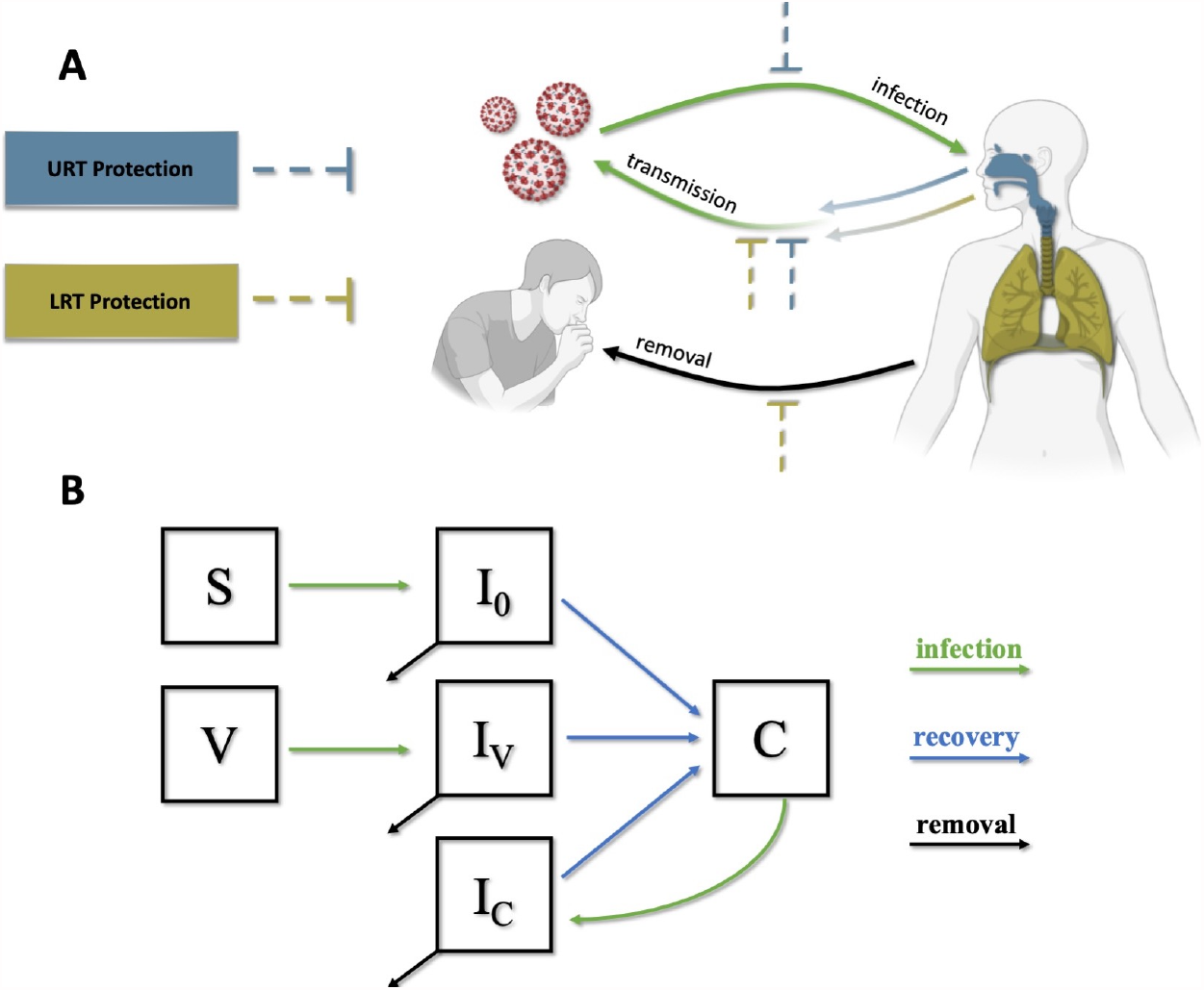
Model Schematic. **(A)** The effects of immunological protection (either vaccinal or naturally acquired) in the URT and LRT on transmission, infection, and disease severity. Immunological protection in the URT and/or LRT decreases the rate of onward transmission. Additionally, URT protection decreases the rate of infection, and LRT protection decreases virulence which we assume to be proportional to the rate of removal from the transmission pool. **(B)** Structure of the epidemiological model. Susceptible (S) and vaccinated individuals (V) become infected (to classes I_0_ and I_V_ respectively) in a density dependent manner and eventually recover to a convalescent class (C) with naturally acquired immunity. These convalescent individuals can become reinfected and return to an infected class (I_C_). All infected classes contribute to transmission.

Our analyses reveal several important relationships between vaccinal immunity and shifts in selection for increased virulence. Most importantly, we observe that selection for increased virulence decreases as vaccinal URT protection increases, and that vaccines generate the most selection for increased virulence when they confer a high degree of LRT protection and a low degree of URT protection (Fig. 3). Additionally, for any given combination of vaccine effects (*r*_*U,V*_, *r*_*L,V*_), selection for increased virulence increases (or selection against increased virulence decreases) with vaccine coverage. Unsurprisingly, we find that increased virulence is always selected for when *α*_*optium*_ > *α*_*B*.1.1.7_, but increased virulence can also be favored when *α*_*optium*_ ≤ *α*_*B*.1.1.7_ (Fig. S1).

**Figure 3:**
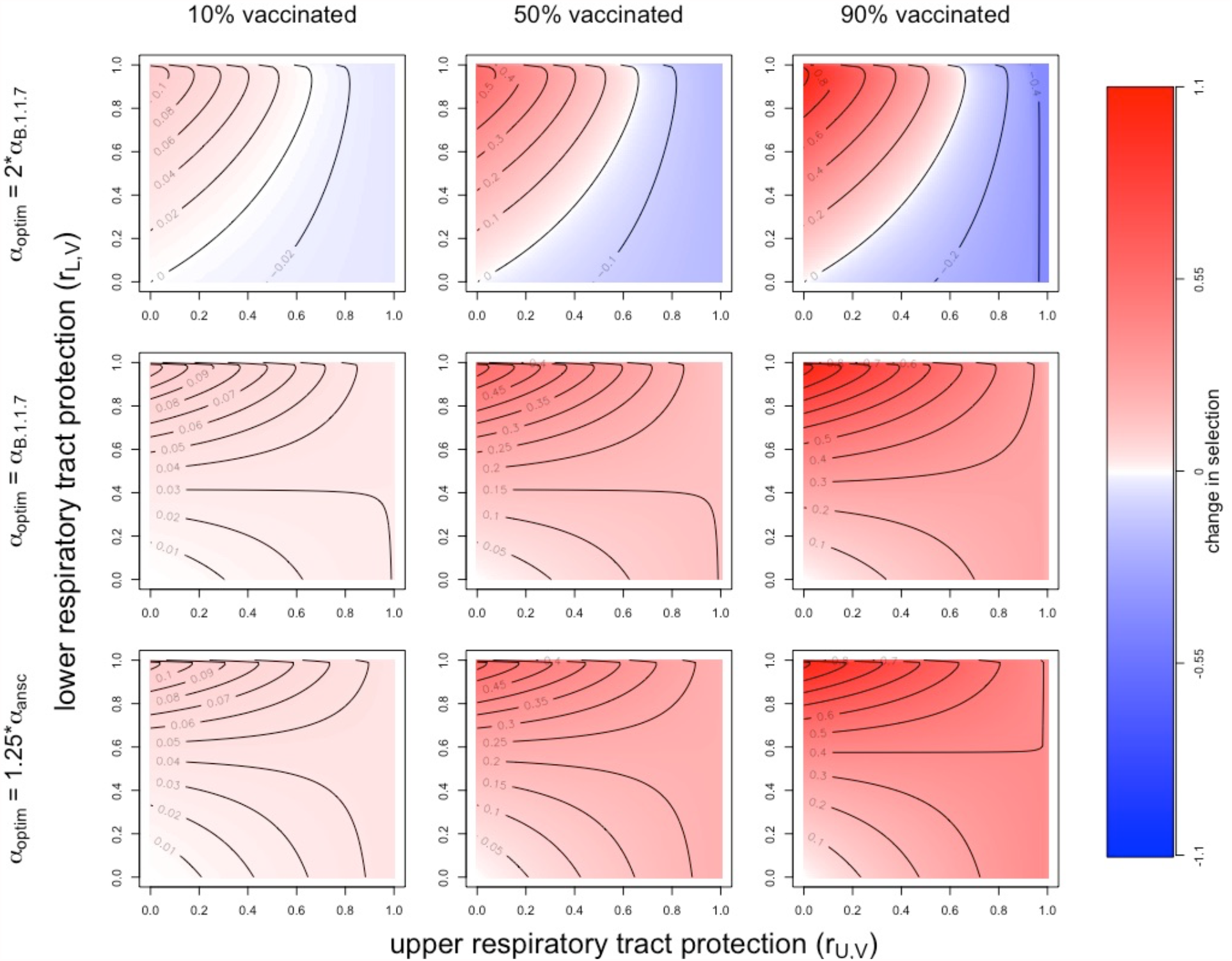
Change in selection for increased virulence associated with vaccinal immunity. Panels show the change in the selection coefficient for an increase in virulence from *α*_*B*.1.1.7_ to 1.5\**α*_*B*.1.1.7_ associated with vaccinal immunity. Blue signifies a decrease in the selection coefficient, and red an increase in the selection coefficient, with darker colors indicating a greater magnitude of change. Note that the directionality of the change in the selection coefficient does not indicate whether the *R*_*E*_ associated with increased virulence is greater or less than one. In all panels natural immunity effects are set to *r*_*U,C*_ = 0.5, *r*_*L,C*_ = 0.75 and the lower and upper respiratory tracts contribute to transmission (*ε* = 0.5).

Beyond the magnitude and directionality of selection for increased virulence, our analyses connect vaccinal immunity to qualitative epidemiological outcomes, capturing the larger overall benefits of vaccines (Fig. 4). Even if a more virulent strain evolves in response to vaccination, if it can be eliminated by vaccination (i.e., herd immunity can be achieved), then the benefits of vaccination outweigh the costs. Across all assumptions about the optimum degree of virulence, herd immunity (*R*_*E*_ < 1) is only achieved when URT protection is strong and vaccine coverage is high. Within the parameter spaces corresponding to herd immunity, increasing URT protection is again associated with better outcomes, first eliminating the potential for the evolution of a more virulent (and more transmissible) strain to bring *R*_*E*_ back above 1, and if *α*_*optium*_ > *α*_*B*.1.1.7_, ultimately negating selection for a variant with a 50% increase in virulence. In the absence of herd immunity, vaccinal immunity can only lead to selection against increased virulence when *α*_*optium*_ = *α*_*B*.1.1.7_. We observed similar patterns of selection for increased virulence and epidemiological outcomes when assuming that LRT infection contributes minimally to transmission and for alternate assumptions about the strength of the protective effects of natural immunity in the URT and LRT (Figs. S2-7).

**Figure 4:**
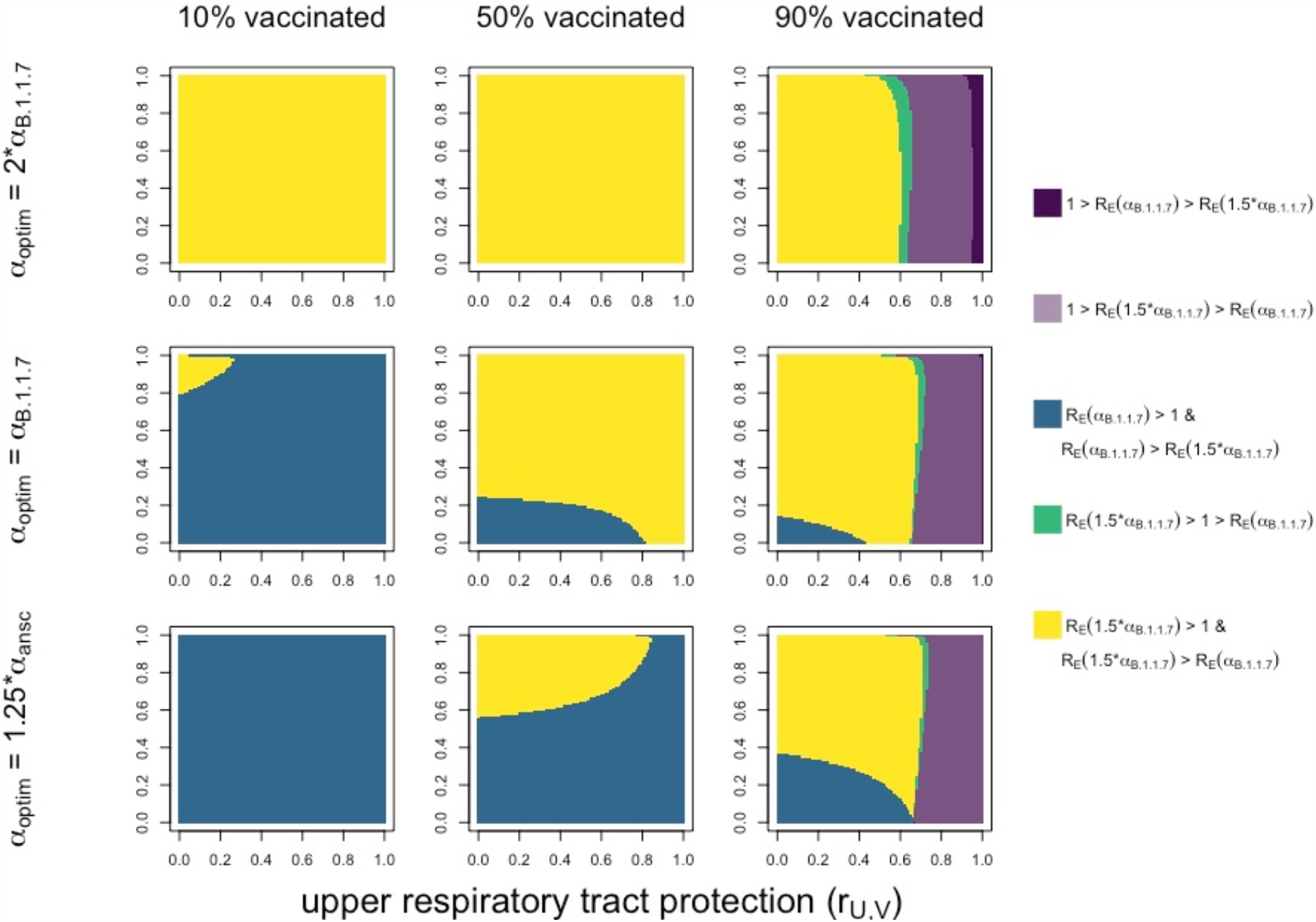
Epidemiological outcomes associated with vaccinal immunity. We assessed the outcomes associated with vaccinal immunity by comparing the R_E_ value associated with observed virulence, *R*_*E*_(*α*_*B*.1.1.7_), to the R_E_ value associated with a 50% increase in virulence, *R*_*E*_(*α*_*B*.1.1.7_ * 1.5). Panels show outcomes associated with different assumptions about optimal virulence and vaccine coverage. Yellow and blue colors indicate selection for and against increased virulence respectively, without vaccinal immunity leading to herd immunity (i.e. *R*_*E*_ > 1). Green indicates that vaccinal immunity induces herd immunity, but that a variant with increased virulence (and transmission) would be selected for and could erode herd immunity. Purple colors indicate that vaccinal immunity leads to herd immunity that cannot be eroded with the introduction of a variant with doubled virulence, regardless of whether selection would act against that variant (dark purple) or in favor of that variant (light purple). In the latter case, the relative frequency of the more virulent variant could increase even though the incidence of infection falls due to herd immunity. Qualitatively similar results were obtained for different assumptions about the contribution LRT infection to transmission (Fig. S3) and the strength of natural immunity (Figs S5, S7).

Comparing current evidence about the effects of COVID-19 vaccines from phase 3 clinical trials to our model predictions can shed light on how we might expect virulence evolution to unfold. Current evidence suggests that approved COVID-19 vaccines provide robust protection against disease. Two mRNA vaccines, BNT162b2 and mRNA-1723, were shown to be 95% and 94% effective in preventing COVID-19 respectively while also exhibiting promise of preventing severe symptoms (3, 4). A recombinant adenovirus vaccine, ChAdOx1 had an efficacy of 62-90% in clinical trials carried out in Brazil, South Africa, and the United Kingdom (5). Another recombinant adenovirus vaccine, Gam-COVID-Vac, or Sputnik V, exhibited an efficacy of 92% (6). The current understanding of infection limiting effects of vaccines is more limited, but nonetheless promising. The ChAdOx1 vaccine was shown to reduce the incidence of PCR confirmed infection by 67% (39), consistent with a moderately strong infection blocking effect.. An even more striking result was observed for the BNT162b2 vaccine, which has been shown to be 95.3% effective in preventing infections (40). Data on the extent to which SARS-COV-2 vaccines reduce onwards transmission is currently lacking. As a whole, this evidence suggests that COVID-19 vaccines confer a very high degree of LRT protection, and likely a significant amount of URT protection, although it is not possible at this time to map vaccines to a specific combination of protective effects (*r*_*U,V*_, *r*_*L,V*_), and the emergent of vaccine-resistant may decrease vaccine efficacy (41). Given this combination of effects, our model suggests that 1) the directionality of potential selection for increased virulence generated by vaccines is highly sensitive to the protective effects of vaccination and the optimum degree of virulence, and thus impossible to identify at this time 2) the strength of this selection for or against increased virulence will increase with vaccine coverage, and 3) existing vaccines will likely significantly reduce transmission and could, if administered to a sufficient proportion of the population, generate herd immunity even if virulence evolution occurs.

Two sets of qualitative outcomes corresponding to changes in the model assumptions are worth noting. First, if a virulence-transmission trade-off does not exist (e.g. transmission increases linearly with virulence), selection for increased virulence will always occur, and the only evolutionary effects of vaccination are to modulate the strength of selection. Second, if increased virulence leads to faster symptom onset and/or a lower asymptomatic rate, the costs of virulence would increase, leading to better outcomes than those predicted in the ‘pessimistic’ scenario presented here, but a model with a different underlying epidemiological structure would be required to make precise predictions.

## Discussion

As the global spread of SARS-CoV-2 continues and the widespread rollout of vaccines is underway, evaluating the risk of vaccine-driven virulence evolution increases in priority. We reviewed the theoretical drivers of virulence evolution, distilled a set of conditions necessary for the evolution of increased virulence, and evaluated them in the context of SARS-CoV-2, concluding that further evolution of SARS-CoV-2 virulence is plausible, but not certain. While the limits to SARS-CoV-2 evolution are not clear, several mechanisms could potentially result in vaccines driving selection for or against increased virulence. Using an evo-epidemiological model, we determined that under a pessimistic set of assumptions, existing vaccines could potentially select for increased virulence, but that if this were to occur, they would still generate herd immunity if administered to a large percentage of the population.

The large degree of uncertainty surrounding the plausibility of vaccine driven virulence evolution, combined with our finding that vaccines could lead to herd immunity even if they select for increased virulence, suggests no cause for immediate concern about the risks posed by vaccines in regard to virulence evolution. However, these factors do point to several actions that should be taken to mitigate any risks. First, the infection blocking and transmission reducing characteristics of all SARS-CoV-2 vaccines should be evaluated, and the use of any vaccine that does not significantly reduce contagion should be carefully considered. Second, as more vaccines and more data become available, governments should prioritize the use of vaccines that provide robust protection against both disease and transmission. Third, vaccination programs should aim to immunize a large percentage of the population as quickly as possible, as herd immunity, which could mitigate selection for increased virulence, is only possible at high vaccine coverage (in the absence of non-pharmaceutical interventions).

The benefits of transmission/infection reducing immunity stemming from vaccinal URT protection extend beyond minimizing selection for increased virulence and generating herd immunity. These effects, along with other measures, such as non-pharmaceutical interventions, would also decrease effective viral population size, diminishing the impact of selection, the genetic diversity of the virus, and thus the likelihood of vaccine driven virulence evolution. However, sources of genetic diversity could exacerbate risks of viral evolution. Significant viral diversity can be generated in chronically infected immunocompromised patients, and enhanced infection control procedures have been recommended to limit the spread of spread of mutations from these individuals. Animal reservoirs of are another potential source of genetic diversity (42). SARS-CoV-2 epidemics in mink farms have already been linked to the generation of novel mutations (43) and animal to human transmission (44). Other domestic and agricultural animal populations, especially those maintained at high density, should be actively monitored for SARS-CoV-2 transmission. Wild animal populations also have potential to harbor sustained viral transmission but present significant challenges for implementing control measures. Species that are known to be competent hosts for coronaviruses and regularly come into contact with humans should be prioritized for monitoring.

If vaccines were to drive the evolution of increased virulence, protection against disease in vaccinated individuals may or may not erode, but other groups would bear greater impacts (27). Unvaccinated individuals would bear the consequences of increased virulence without any vaccinal protection to offset disease severity. Vaccine driven evolution could also amplify healthcare disparities between populations if a SARS-CoV-2 strain with increased virulence evolves in a vaccinated population, and spills over into an unvaccinated population. Beyond seeking to reduce disparities in vaccination, countries with the highest vaccine coverage should all engage in virulence monitoring to detect emergent variants with increased virulence, and if such variants arise, enact containment measures to mitigate negative consequences for countries with limited or no access to vaccines.

While our analyses give approximate expectations under a specific set of assumptions, robust expectations about the likelihood, magnitude, and trajectory of virulence evolution remain elusive. We did not include immunological waning in our model, but the loss of vaccinal and/or natural immunity over time could increase the both effective viral population sizes and likelihood of SARS-CoV-2 becoming endemic (45–47). Future analyses should explore how waning immunity might alter selection for increased virulence as evidence regarding the longevity of immunity emerges. Our theoretical analysis assumed a conventional saturating positive relationship between transmission and virulence. However, the shape, nature, and existence of any trade-offs governing virulence evolution in SARS-CoV-2 are currently unknown, and the form of true relationship could modify the direction and magnitude of selection acting on virulence. Assessments of the risk of virulence evolution should be updated as new findings emerge, both by directly monitoring the virulence of the virus, and by better characterizing the nature of the tradeoff. To monitor virulence, infection fatality ratios associated with circulating SARS-CoV-2 strains should be surveilled to detect early evidence of any evolutionary trends. Stratifying these observations by age and immunological status (naïve, natural immunity, vaccinal immunity) would allow for changes in population-wide infection fatality rates to be properly attributed changes in virulence or other factors. Decreases in fatality rates tied to medical advances will complicate this monitoring, especially as they vary globally, but animal or cell-culture models might provide a means for standardized measurement. Trade-offs affecting the evolution of SARS-CoV-2 transmission can be characterized through comparative epidemiological analyses and clinical studies of within-host viral dynamics.

While we focus on the evolution of quantitative virulence in this paper, other aspects of viral evolution should also be considered when evaluating candidate SARS-CoV-2 vaccines. Antigenic evolution leading to ‘vaccine escape’ (i.e. resistance) has occurred in response to other human vaccines (48, 49) and the potential for circulating SARS-CoV-2 variants to evade vaccinal immunity is a current issue of concern (50). The evolution of resistance is theorized to be more likely to occur in response to drug treatment than vaccination because drugs often target a single pathogen epitope while vaccines induce an immunological response with a border target set (51). All of the candidate SARS-CoV-2 vaccines that have completed phase 3 trials induce an immune response targeted only at the viral spike protein (31), and as such could be viewed as having effects intermediate between drugs and other vaccines. However, the necessity of the spike protein for cellular entry (52) might make SARS-CoV-2 vaccines unlikely to drive the evolution of resistance relative to other vaccines that include many proteins. While evidence thus far suggests that low viral genetic diversity might limit the evolution of vaccine escape (53), the potential for this event to occur should be assessed by comparing patterns of within-host viral evolution between placebo and vaccine groups in ongoing clinical trials (7). Antigenic evolution may also have implications for virulence evolution. This could occur through either direct effects, such as antigenic changes altering the function of proteins involved in transmission or pathogenesis, or through indirect effects, such as antibody dependent enhancement (54), which could lead to increases in viral load and ultimately greater disease severity.

Vaccines have the ability to provide protection against COVID-19 disease at a massive scale, but widespread protection could impose significant selection pressures on the SARS-CoV-2 virus. Current evidence suggests no immediate cause for concern, but careful monitoring and efficacy evaluation are needed moving forward.

## Methods

### Epidemiological Model

To evaluate the potential evolutionary consequences associated with SARS-CoV-2 vaccine candidates, we develop a compartmental ordinary differential equation model that broadly reflects SARS-CoV-2 epidemiology and the protective effects of naturally acquired and vaccine-induced immunity (Fig. 2). We extend previous work by explicitly separating the impacts of vaccinal and natural immunity in the upper and lower respiratory tracts, and the impact of these protective effects on infection, transmission, and virulence. We assume that infection in the LRT and URT additively contributes to transmission rate, *β* (Eqn. 1). The parameter *ε* defines the fractional contribution of the LRT to transmission. Transmission rates in both the URT and LRT follow an increasing and saturating function of virulence defined by parameters b_1_ and b_2_. We assume that a virulence rate greater than 0.0025 is necessary for transmission (Eqn. 1). Vaccine-induced immunity limits transmission by reducing virulence by a scalar 1 − *r*_*U,V*_ in the upper respiratory system, and 1 − *r*_*L,V*_ in the lower respiratory tracts (in this case transmission is calculated as *β*(*α, r*_*U,V*,_ *r*_*L,V*_)). Likewise, naturally acquired immunity limits transmission by reducing virulence by a scalar of 1 − *r*_*U,C*_ in the URT and 1 − *r*_*L,C*_ in the LRT (in this case transmission is calculated as *β*(*α, r*_*U,C*,_ *r*_*L,C*_)).

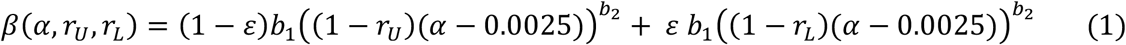

Moving from how immunity affects transmission to considering how it might affect infection and virulence, we begin by connecting virulence to the rate of removal of infectious individuals from the transmission pool. We set this rate equal to virulence (*α*) times a constant (*p*). This association between virulence and the removal of infectious individuals combined with the form of the function relating virulence to transmission constitutes a virulence-transmission trade-off (Fig. 1). To incorporate the effects of immunity, we assume that only LRT protection reduces virulence, in line with current reporting on the pathology of severe cases of SARS-CoV-2. The effects of virulence on disease associated removal are reduced by a scalar (1 − *r*_*L,V*_) for vaccinated individuals and by (1 − *r*_*L,C*_) for individuals with naturally acquired immunity. To model effects of immunity on transmission, we assume that the force of infection (*λ*) experienced by vaccinated individuals is reduced by a scalar (1 − *r*_*U,V*_) for vaccinated individuals and by (1 − *r*_*U,C*_) for individuals with naturally acquired immunity, as the URT seems to be the key driver of transmission.

This framework for modeling the effects of immunity is designed to capture key differences between URT and LRT protection. To consider how vaccinal and natural immunity affect infection, transmission, and virulence without separating the effects of immunity between respiratory compartments, this framework would need to be adjusted to decouple reductions in transmission from reductions in infection and virulence.

After defining the relationships between immunity, transmission, infection, and virulence, we incorporate them into a compartmental epidemiological model (Fig. 2B, Eqn. 1-3). We assume a one day time step, and accordingly set the recovery rate *γ* = 1/7 (55). We ignore disease-associated mortality, assuming that isolation occurs before death. This is consistent with the observation that death usually occurs several weeks after the appearance of symptoms. As this model is aimed at making short term predictions about the effects of vaccination on selection for virulence, we ignore births, non-COVID-19 associated deaths, and the waning of immunity. Note that convalescent individuals can become reinfected in the absence of immunological waning if immunity provides incomplete protection against reinfection (*r*_*U,C*_ < 1).

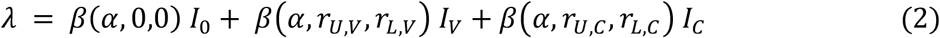

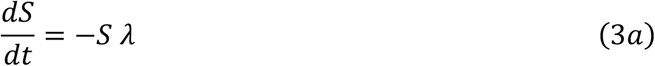

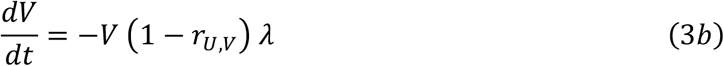

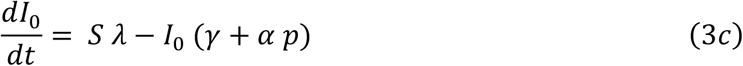

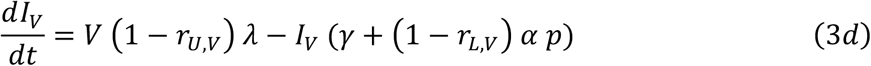

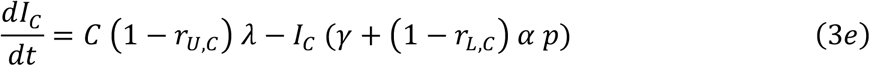

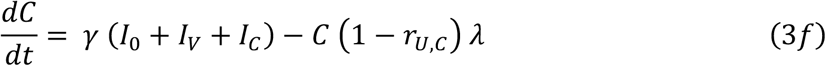

The dynamics of this model hinge on the relationship between transmission and virulence, and as such are sensitive to the shape of the virulence-transmission trade-off. We aim explore how the evolutionary dynamics predicted by this model change with assumptions about where the virulence of *B*.*1*.*1*.*7* falls on the trade-off curve. We begin by defining the observed values of R_0_ 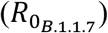 and virulence (*α*_*B*.1.1.7_) associated with this strain. As *B*.*1*.*1*.*7* has been linked to an approximately 50% increase in case fatality rate (56, 57), and the infection fatality rate of the ancestral strain (*α*_*ansc*_) is estimated to be 0.005 (58), we assume *α*_*B*.1.1.7_ = 1.5 * *α*_*ansc*_ = 0.0075. Likewise, the transmission rate of *B*.*1*.*1*.*7* is estimated to be approximately 1.5 times greater than that of the ancestral strain. Given the basic reproduction number of the ancestral strain 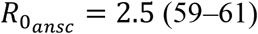, we set 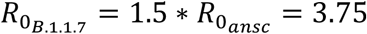 Following this, we set *p* = 66.666 so that the average amount of time an individual is infectious before isolation is, 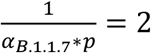 days if recovery is ignored. This value is roughly consistent with the assumptions of other models (62). Next, we construct a set of optimal virulence (*α*_*optium*_) values to explore that reflects various assumptions about the fitness of *α*_*B*.1.1.7_ and *α*_*ansc*_relative the optimum. Because the *B*.*1*.*1*.*7* strain is assumed to be more fit than the ancestral strain, we assume that the optimum virulence is either intermediate between *α*_*ansc*_ and *α*_*B*.1.1.7_, or greater than and *α*_*B*.1.1.7_: *α*_*optium*_ ∈ {1.25 * *α*_*ansc*_, *α*_*B*.1.1.7,_ 2.0 * *α*_*B*.1.1.7_}. For a given assumption about the value of *α*_*optium*_, we use the next-generation matrix approach (63) to set the value of b_2_ to that which maximizes R_0_ at *α*_*optium*_ and b_1_=1. By definition, R_0_ is calculated assuming a completely susceptible population. Next, we used these same methods to set the value of b_1_ to that which corresponded 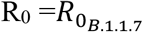 at *α*_*B*.1.1.7_. We checked to ensure that the value of b_2_ was insensitive to the value of b_1_, that *α*_*optium*_ maximized R_0_, and that *α*_*B*.1.1.7_ correspond to 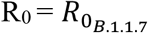 at the identified values of b_1_ and b_2_. Figure 1 shows the virulence transmission trade-off curves constructed from the three assumptions about the value of *α*_*optium*_; these assumptions map to a wide range of possible shapes of the trade-off.

### Evolutionary Analysis

To characterize the landscape of short-term selection on pathogen virulence in response to vaccination across a range of characteristics of vaccinal protection in the URT and LRT, we identify the strength of selection (fitness differential) for a ‘next step’ towards increased virulence, which for the purposes of our analysis we define as a 50% increase in virulence relative to *α*_*B*.1.1.7_. While more formal methods exist for analyzing the trajectory of virulence evolution in the long and short term, the current lack of knowledge surrounding relevant aspects of SARS-CoV-2 biology make these approaches less suitable for characterizing the approximate directionality of short term virulence evolution in current data-sparse circumstances. The adaptive dynamics toolkit provides a means to characterize the trajectory of virulence evolution in the long term as viral evolution perturbs epidemiological equilibria (64). Over this time scale, waning immunity will likely emerge as a key factor driving the epidemiology of COVID-19 (47). An adaptive dynamics approach would require strong assumptions about the duration of immunity that would extend beyond the scope of existing data. Once appropriate data are available, this type of an analysis can be used to generate robust predictions. Approaches linking viral mutation to epidemiological dynamics can be used to characterize the trajectory of virulence evolution over shorter epidemiological time scales, but these methods require strong assumptions about the mutation rates and the probability of mutations altering virulence (65). Given that the links between SARS-CoV-2 mutations and virulence remain largely uncharacterized, and that a large proportion of mutations may occur in rare events involving chronic infections of immunocompromised individuals (42), these methods currently have limited utility. Once paired with suitable evidence, such as a genotype-to-phenotype map for SARS-CoV-2 virulence [Glennon et al. Epidemics], these methods could generate deep insights.

We initialize the model with 25% of individuals convalescent (roughly consistent with the estimated cumulative incidence in the U.S. as of December 2020 (66)), and all non-vaccinated individuals susceptible. For a given set of protective effects (*r*_*U,V*,_ *r*_*L,V*_), we calculated R_E_ for *α*_*B*.1.1.7_ using the next-generation matrix approach. Next, we used the same methods to calculate R_E_ for *α* = 1.5 * *α*_*B*.1.1.7_. We then calculated the selection coefficient for increased virulence as the difference between R_E_ at 1.5* *α*_*B*.1.1.7_ and R_E_ at *α*_*B*.1.1.7_.

## Data Availability

All code used to perform the analyses presented in this manuscript is available on GitHub.

https://github.com/ianfmiller/covid_vaccines_virulence_evolution

## General

We thank Bryan Grenfell, Jeremy Farrar, Gordon Douglas, Daniel Douek, and Adrian McDermott for helpful discussions. Figure 2A was created in part using BioRender.com.

## Author contributions

IFM and CJEM conceived of the study. IFM and CJEM reviewed the literature, and IFM performed the analyses. IFM and CJEM wrote and revised the manuscript.

## Data and materials availability

All code used in the analyses is available on GitHub (67).

## Supporting Information

Figures S1-S7

## Supporting Information

**Figure S1:**
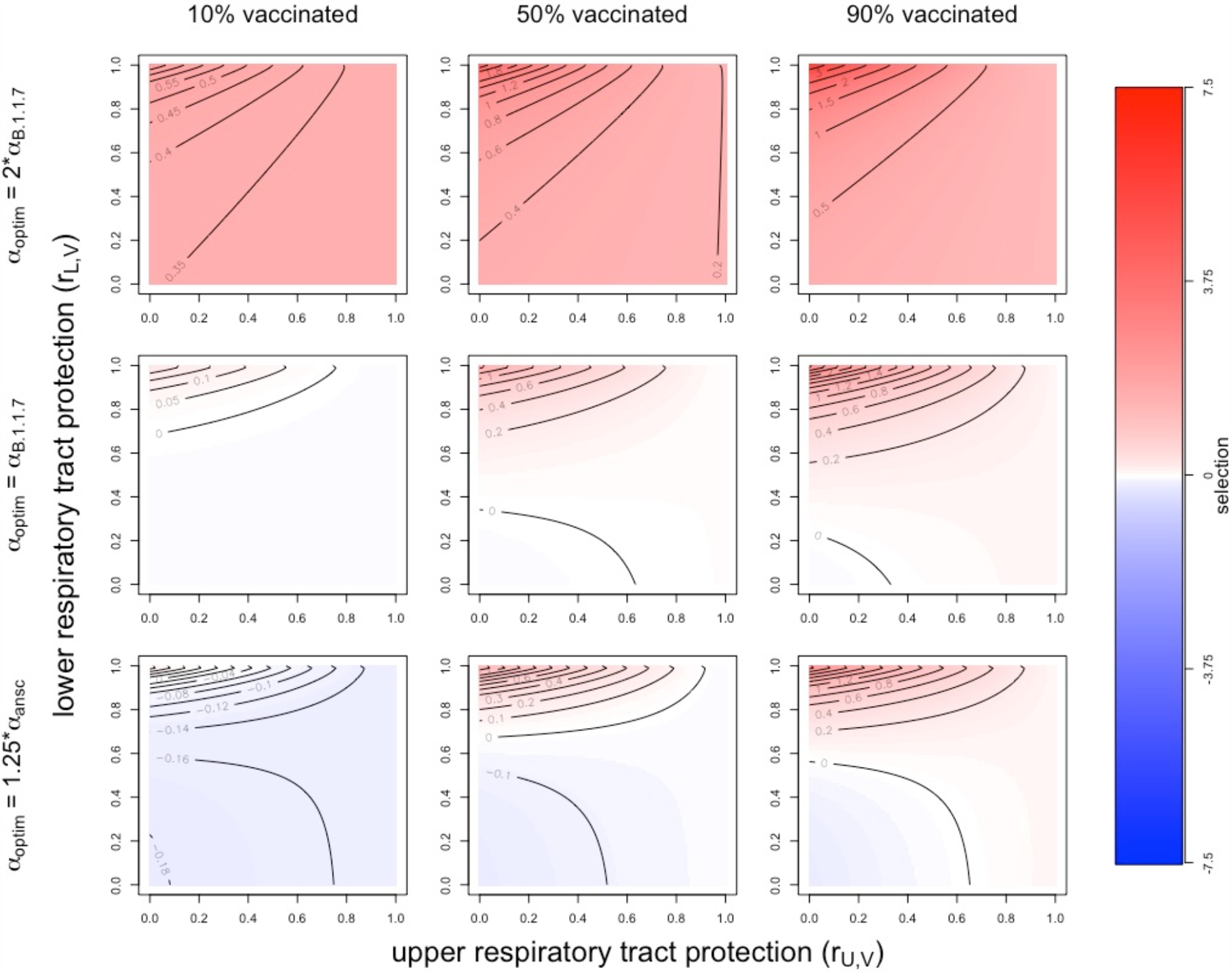
Vaccinal immunity and selection for and against increased virulence. Panels show the strength and direction of selection for an increase in virulence from *α*_*B*.1.1.7_ to *α*_*B*.1.1.7_ * across a range of assumptions about optimal virulence and vaccine coverage. Blue signifies selection against increased virulence, and red signifies selection for increased virulence, with darker colors indicating stronger selection. In all panels natural immunity effects are set to *r*_*U,C*_ = 0.5, *r*_*L,C*_ = 0.75 and the lower and upper respiratory tracts contribute to transmission (*ε* = 0.5). Qualitatively similar results were obtained for different assumptions about the contribution LRT infection to transmission (Fig. S2) and the strength of natural immunity (Figs S4, S6).

**Figure S2:**
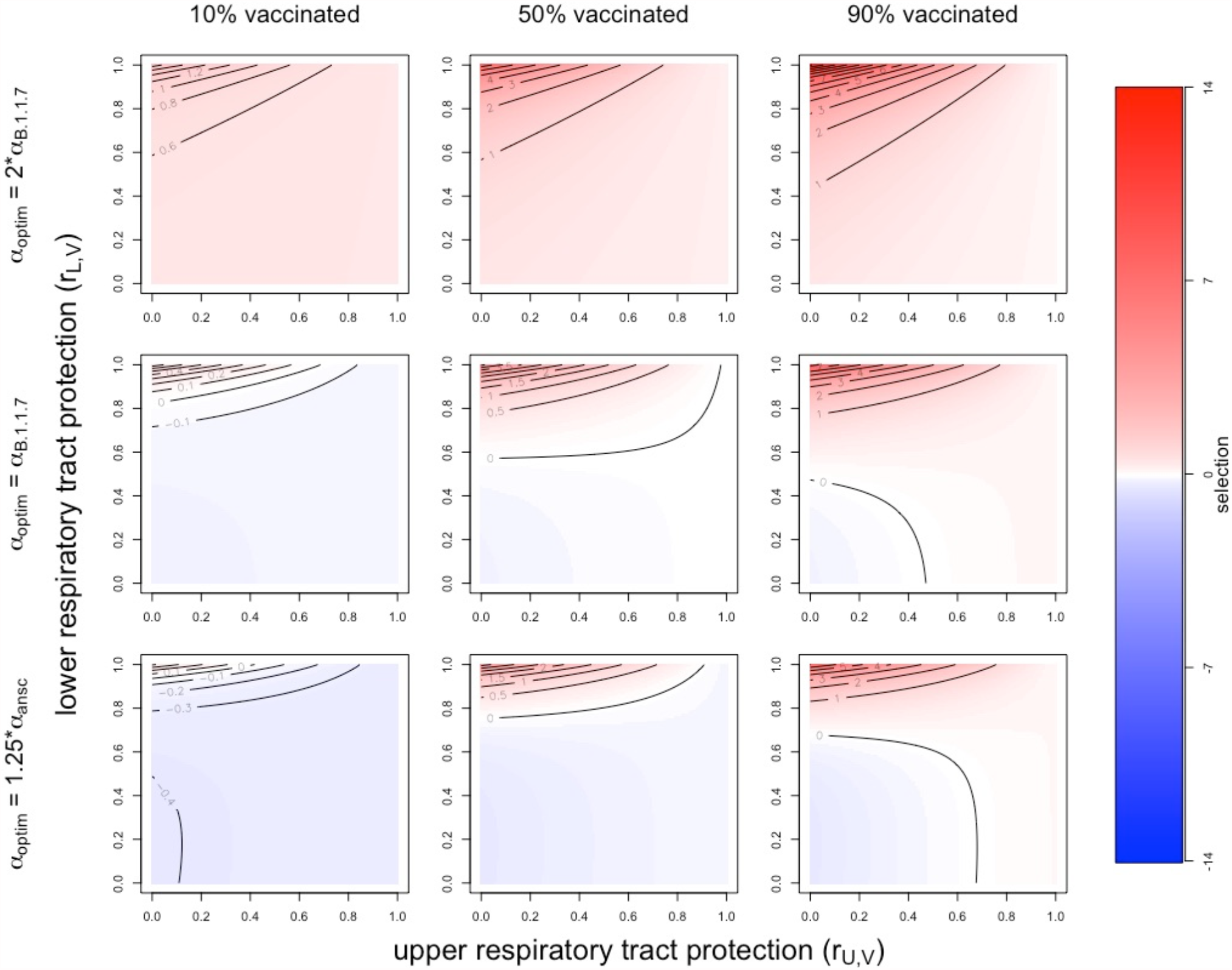
Vaccinal immunity and selection for and against increased virulence when LRT infection contributes minimally to transmission. We found that patterns of selection for increased virulence are qualitatively similar when infection in the URT and LRT contribute equally to transmission (*ε* = 0.5, Fig. 3), and when LRT infection contributes minimally to transmission (*ε* = 0.1, this figure). Panels show the strength and direction of selection for an increase in virulence from *α*_*B*.1.1.7_ to 1.5\**α*_*B*.1.1.7_ across a range of assumptions about optimal virulence and vaccine coverage. In all panels *r*_*U,C*_ = 0.5, *r*_*L,C*_ = 0.75.

**Figure S3:**
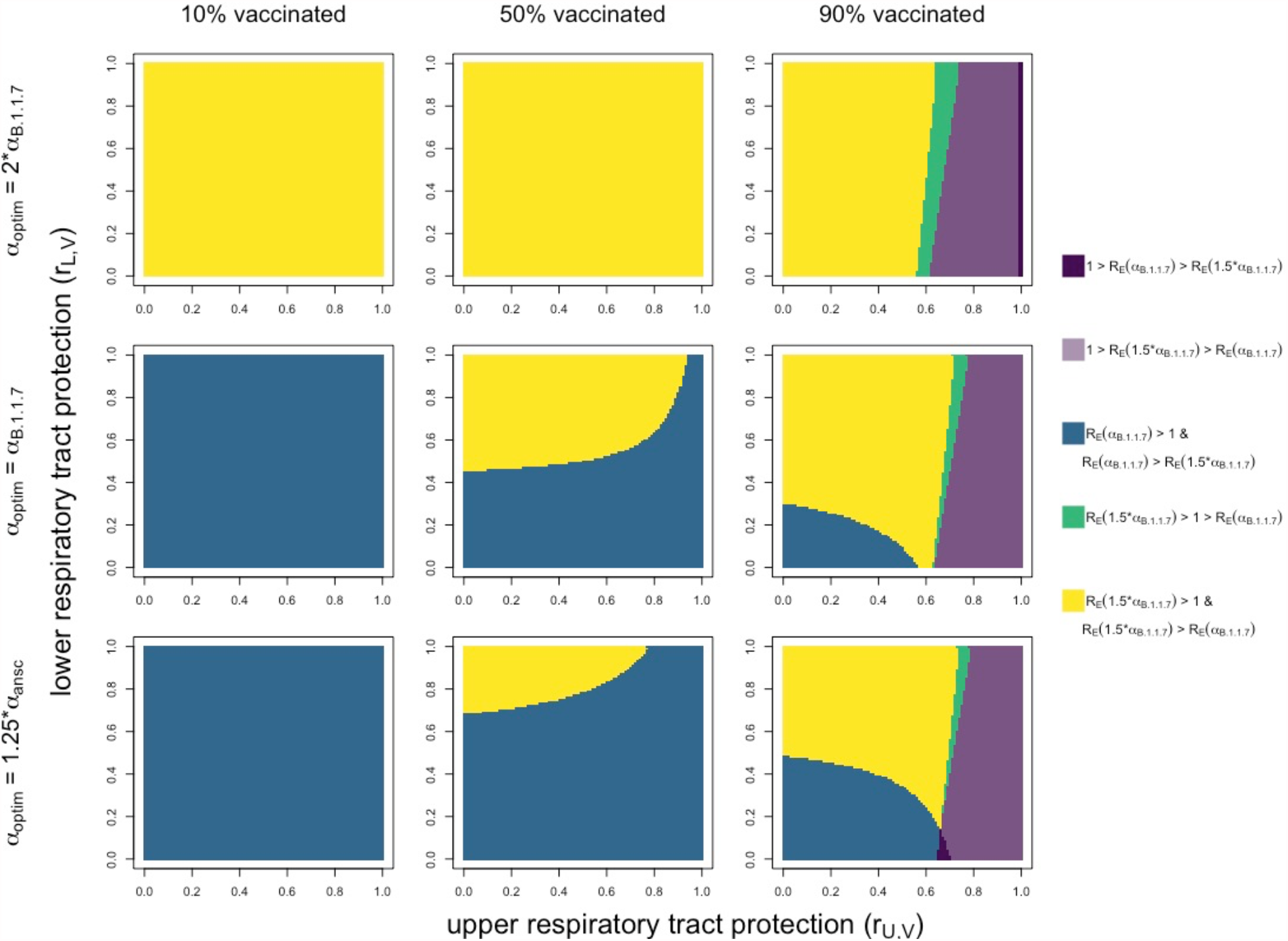
Epidemiological outcomes associated with vaccinal immunity when LRT infection contributes minimally to transmission. The outcomes associated with vaccinal immunity are qualitatively similar when infection in the URT and LRT contribute equally to transmission (*ε* = 0.5, Fig. 4), and when LRT infection contributes minimally to transmission (*ε* = 0.1, this figure). Panels show outcomes associated with different assumptions about optimal virulence and vaccine coverage. In all panels *r*_*U,C*_ = 0.5, *r*_*L,C*_ = 0.75.

**Figure S4:**
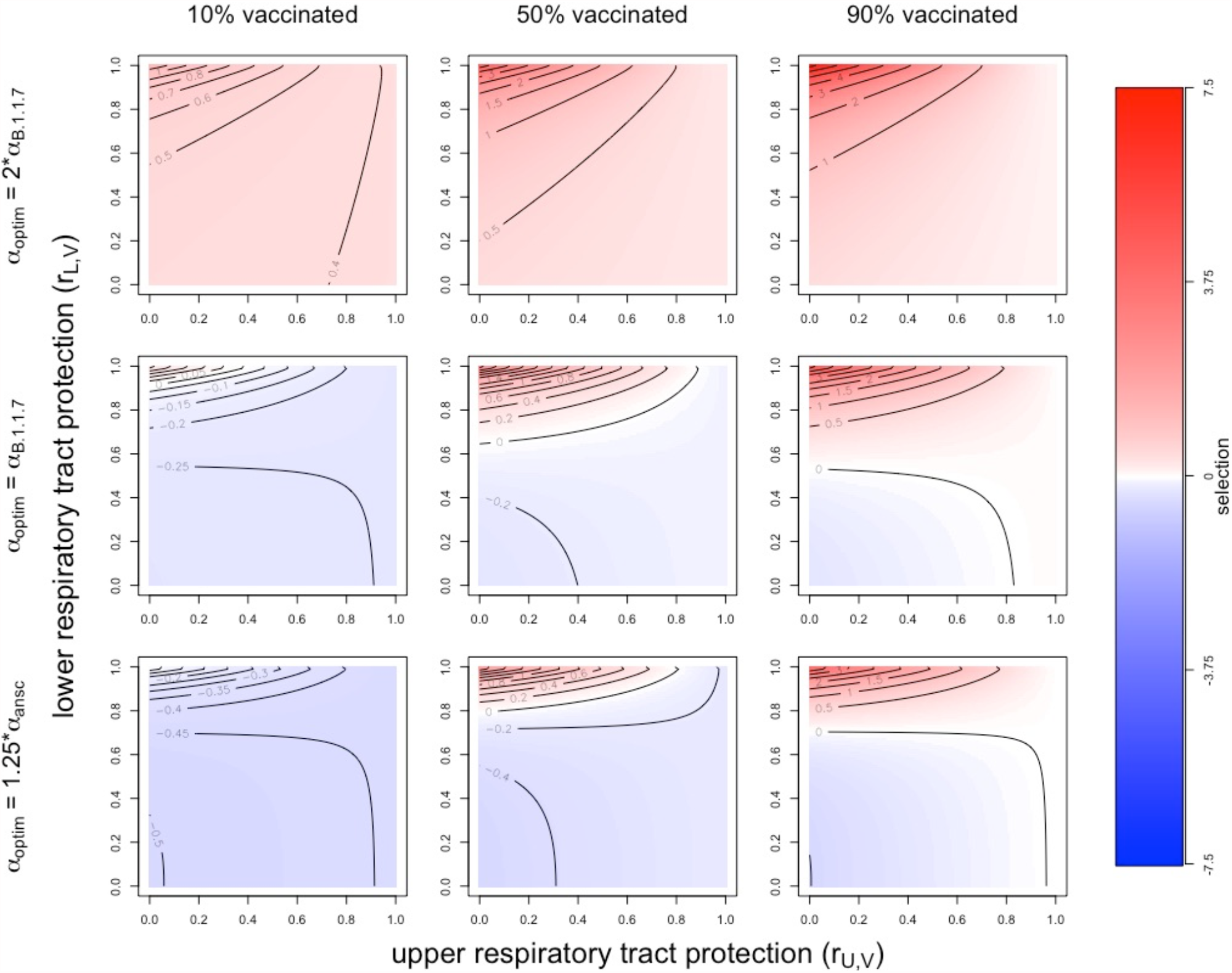
Vaccinal immunity and selection for and against increased virulence with weak effects of natural immunity. Our analyses returned qualitatively similar results across a range of assumptions about the strength of natural immunity. This figure shows results for weaker assumed effects of immunity (*r*_*U,C*_ = 0.25, *r*_*L,C*_ = 0.5) than in the analyses presented in the main text (*r*_*U,C*_ = 0.5, *r*_*L,C*_ = 0.75, Fig. 3). Panels show the strength and direction of selection for an increase in virulence from *α*_*B*.1.1.7_ to 1.5\**α*_*B*.1.1.7_ across a range of assumptions about optimal virulence and vaccine coverage. The lower and upper respiratory tracts contribute equally to transmission (*ε* = 0.5).

**Figure S5:**
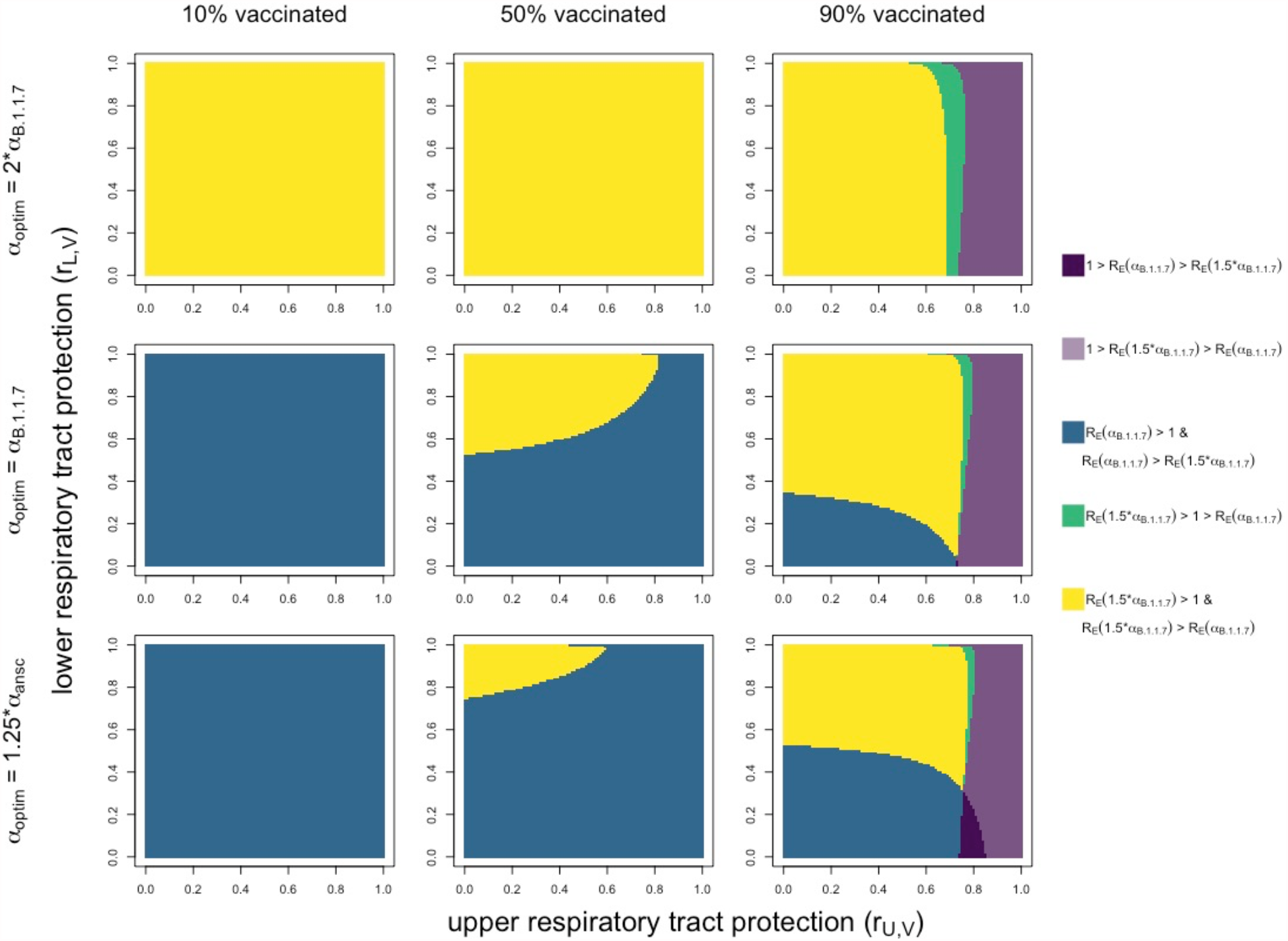
Epidemiological outcomes associated with vaccinal immunity with weak effects of natural immunity. The epidemiological outcomes associated with vaccinal immunity were qualitatively similar across a range of assumptions about the strength of natural immunity. This figure shows results for weaker assumed effects of immunity (*r*_*U,C*_ = 0.25, *r*_*L,C*_ = 0.5) than in the analyses presented in the main text (*r*_*U,C*_ = 0.5, *r*_*L,C*_ = 0.75, Fig. 4). Panels show outcomes associated with different assumptions about optimal virulence and vaccine coverage. The lower and upper respiratory tracts contribute equally to transmission (*ε* = 0.5).

**Figure S6:**
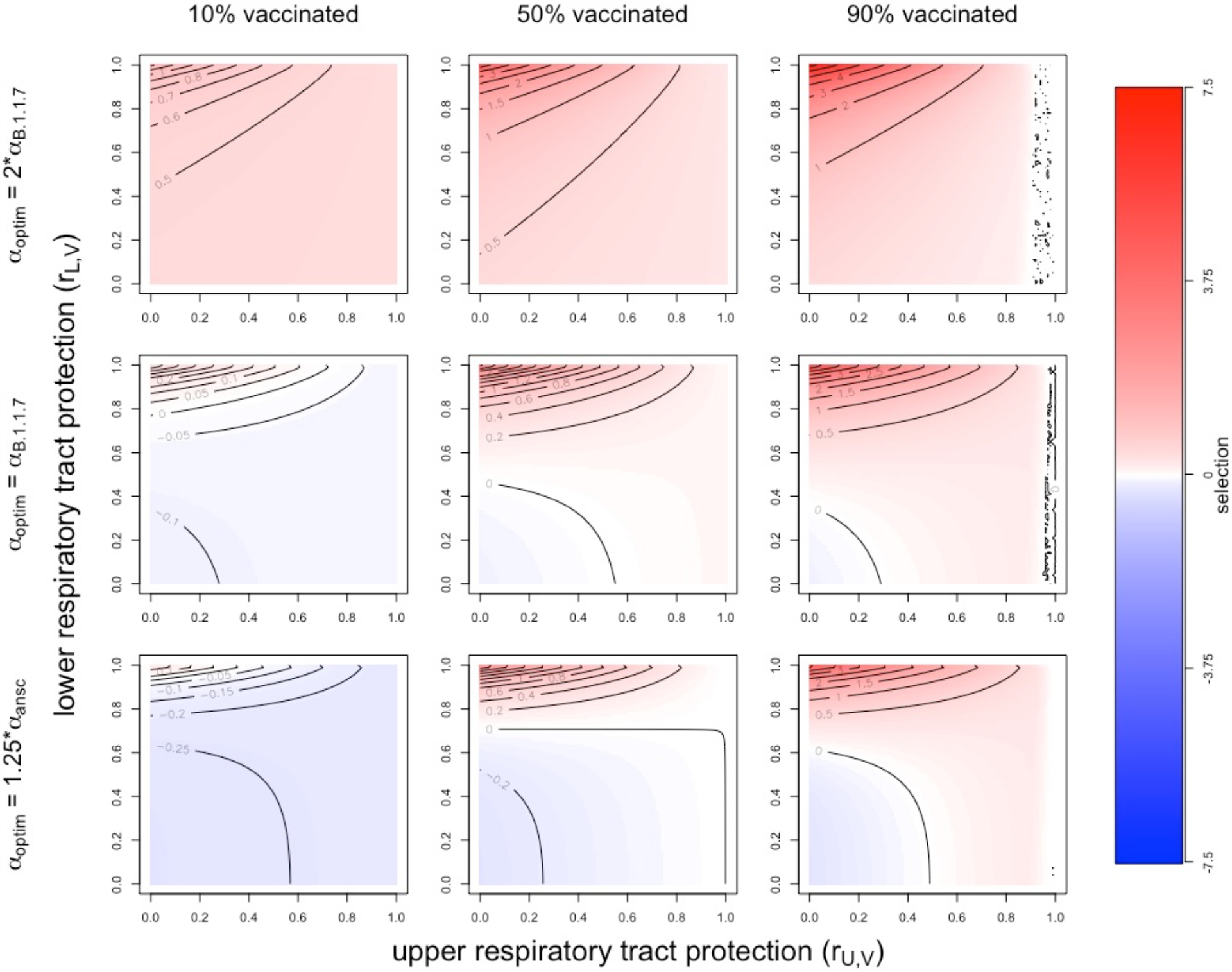
Vaccinal immunity and selection for and against increased virulence with strong effects of natural immunity. Our analyses returned qualitatively similar results across a range of assumptions about the strength of natural immunity. This figure shows results for stronger assumed effects of immunity (*r*_*U,C*_ = 0.75, *r*_*L,C*_ = 1.0) than in the analyses presented in the main text (*r*_*U,C*_ = 0.5, *r*_*L,C*_ = 0.75, Fig. 3). Panels show the strength and direction of selection for an increase in virulence from *α*_*B*.1.1.7_ to 1.5\**α*_*B*.1.1.7_ across a range of assumptions about optimal virulence and vaccine coverage. The complex contour patterns on the far right of panels showing results for 90% vaccine correspond to a selection coefficient of 0. The lower and upper respiratory tracts equally contribute to transmission (*ε* = 0.5).

**Figure S7:**
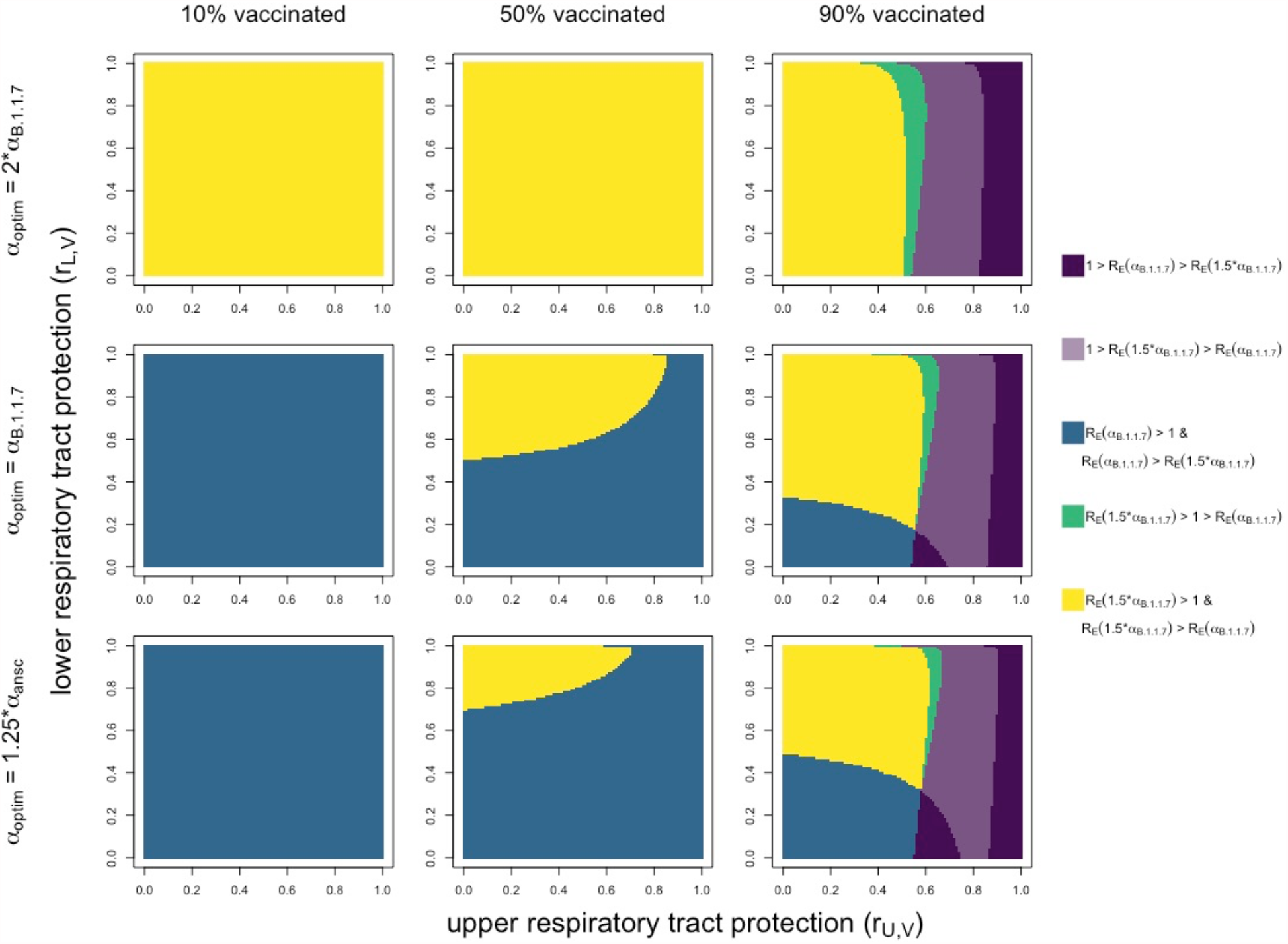
Epidemiological outcomes associated with vaccinal immunity with strong effects of natural immunity. The epidemiological outcomes associated with vaccinal immunity were qualitatively similar across a range of assumptions about the strength of natural immunity. This figure shows results for stronger assumed effects of immunity (*r*_*U,C*_ = 0.5, *r*_*L,C*_ = 1.0) than in the analyses presented in the main text (*r*_*U,C*_ = 0.5, *r*_*L,C*_ = 0.75, Fig. 4). Panels show outcomes associated with different assumptions about optimal virulence and vaccine coverage. The lower and upper respiratory tracts contribute to transmission (*ε* = 0.5).

